# Antibiotic utilization pattern in burn patients admitted at tertiary hospital: A retrospective study

**DOI:** 10.1101/2022.02.15.22270999

**Authors:** Prakriti Thapa, Durga Bista, Pankaj Baidya, Piyush Giri

## Abstract

Burn patients are at high risk for nosocomial infection. Antibiotics are the key drugs for the treatment of infections. Overuse and inappropriate use of antibiotics increase both bacterial resistance and the cost of treatment. The introduction of correct and rational use of antibiotics appears to be impossible without having the knowledge of the current situation of antibiotic consumption. So, the study was conducted to know the current situation of antibiotic utilization pattern in burn patients.

**Methods:** A Retrospective review of medical records was done to analyze the utilization pattern of antibiotics. The data were collected from Kirtipur hospital from June 2018 to May 2019. All the admitted patients irrespective of age, gender who were prescribed antibiotics and presented within three days of burn were included in the study. Patients admitted for less than 24hrs of a time were excluded from the study.

**Results:** A total of 249 reviewed case records came under inclusion criteria. Among them 51.8% were female and 48.2% were male. Mostly affected age group was 15-29 years (34.5%). Flame burn (51.8%) was the main cause of the burn. The majority had second-degree burn and 36.90% had 0-10% burn. Third-generation cephalosporin, ceftriaxone had the highest DDD/100BD (19.05). The most frequently used antibiotics were ceftriaxone, cefazolin, and piperacillin+tazobactam. DU90% comprises 12 antibiotics out of 30 antibiotics. The average number of antibiotics prescribed was 2.12 with a range of 1 to 7.

**Conclusions:** This study revealed the trend of antibiotic utilization pattern in burn patients. Third-generation cephalosporin, ceftriaxone was the most prescribed antibiotic. Regular antibiotic consumption using DDD methodology is needed for regular monitoring of antibiotic consumption so that timely intervention can be made and this study can be used as a baseline study.

## Introduction

Burn injury is the third most common injury in rural Nepal after fall injury and road traffic injuries that account for five percent of disabilities [1]. Burn injuries may be due to flame burn, scald burn, electrical burn, chemical burn, contact burn, radiation burn, burn due to lightening and inhalational burn [2]. It results in a serious effect on physical, psychological, and financial status in both victim and their entire family [3].

Burn patients are at high risk for nosocomial infection mainly due to a disruption of the skin barrier and to a dysbiosis of the immune system [2, 3]. Infections impair cicatrization, lead to graft loss, prolong treatment, and therefore, extend the length of stay and also increase costs. The most frequent sites of infection are burn wounds, respiratory tract, urinary tract, and bloodstream [2].

Antibiotics are the key drugs for the treatment of infections and are among the most commonly prescribed drugs. Patients can be benefitted from the right use of antibiotics, inappropriate prescription lead to harmful consequences [4]. Overuse and inappropriate use of antibiotics lead to an increase in both bacterial resistance and cost of treatment [5]. Moreover, nosocomial organisms are commonly seen infecting the burn wounds and have Multidrug-resistant (MDR) antimicrobial profiles. Thus, antibiotic resistance rates and higher healthcare costs resulting from drug-resistant infections can be lowered by reducing the consumption of unnecessary antibiotics. Studies have revealed that incorrect choice of antibiotics or its duration of action persists in 25% to 75% of cases [6]. It is very important to prescribe drug rationally especially in developing countries where the limited fund is available in health care and drugs [7]. Without knowledge of the current situation of antibiotic consumption, the introduction of correct and rational use of antibiotics seems to be impossible [8].

An important aspect of antibiotic use is antibiotic consumption. There are several metrics of measuring antibiotic consumption [6]. One of the methods is developed and maintained by the WHO Collaborating Centre for Drug Statistics Methodology [9]. It uses Anatomical Therapeutic Chemical (ATC) as a classification system and it classifies antibiotics as pharmacologically active substances based on the organ or system on which they act and on their therapeutic, pharmacological, and chemical properties. It uses DDD as a unit of measure. The DDD is the assumed average maintenance dose per day of an antibiotic substance(s) used for its main indication in adults and is only assigned for medicines given an ATC code. When antibiotic consumption is expressed in DDD per 100 patient-days (DDD/100-BD), in-hospital drug use can be analyzed [10]. DU90% measures the number of drugs accounting for 90% of the use in DDDs [11]. The antibiotic utilization study helps in analyzing and evaluating the prescribing patterns of medical professionals and also helps in formulating both drug and antibiotics policy [12].

There is a paucity of such studies on the international level [13]. To the best of my knowledge, no study has been conducted on antibiotic utilization pattern among burn patients in Nepal. This study will give an insight into the antibiotic utilization pattern in burn patients, thus gives reference about their effectiveness and will be helpful as markers for improvement.

## Methods

This study was a retrospective study carried out in Kirtipur Hospital, which is a specialized burn center in Nepal. The medical records of all the patients admitted to the hospital from June 2018 to May 2019 were reviewed. Ethical approval was obtained before data collection from the Kirtipur Hospital (Approval no: 012-2019). The total medical records of 249 patients who met the inclusion criteria were collected from the medical record section of the hospital. All patients with acute burn irrespective of age and gender who were admitted in the hospital, patients receiving antibiotics, the patient who have completed hospital discharge prescription, and patients presented within three days of burn were included in the study. However, patients who were admitted for less than 24 hrs of a time were excluded from the study. Except for pharmaceutical forms like topical and ophthalmological, all other antibiotics given by oral and parental route were included in the study. The age, gender, TBSA, cause of burn and depth of burn, microorganism resistant profile were noted. Also, the name of antibiotics, doses, dosage form, dose frequency, route of administration, duration was recorded. For resistance profile, it was noted either single drug resistance or Multidrug resistance (MDR) or extensively drug resistance (XDR). MDR bacteria were defined as bacteria that are non-susceptible to at least one antimicrobial agent in three or more antimicrobial classes. Extensively drug resistance (XDR) was defined as non-susceptibility to at least one antimicrobial agent in all but two or fewer antimicrobial classes (that is, bacterial isolates remain susceptible to the agent(s) in only one or two classes).

For easier comparison, the amount of antibiotics in gram was converted to the defined daily dose which is a unit based on average daily used for the main indication for the consumption of certain medications.

The following indicators were calculated:

### DDD

of each antibiotic given to the burn patients was calculated. DDD is assumed average maintenance dose per day for a drug used for its main indication in adults.

DDD=Total dose given to patients/DDD

### DDD/100BD

The DDDs per 100 bed days may be applied when drug use by inpatients is considered.

DDD/100BD=Total consumption in DDD/Number of bed days*100

### DU90%

It is calculated by ranking the drug used by volume of defined daily dose and determining how many drugs accounted for 90% of drug use.

The antibiotics like Ciprofloxacin and amoxicillin-clavulanic acid had two DDD. After calculating with their respective DDD, those DDDs were added.

The data were analyzed and tabulated using MS-Excel (version 7).

## Results

The patient that met the inclusion criteria was 249. The total length of stay of included patients was 2694 days. Out of total patients, 129 (51.80%) were females and 120 (48.20%) were males. Out of a total of 249 patients, 86 (34.54%) of the patients belonged to age group 15-29 years, followed by an aged group less than 14, 30 to 44 years, 45 to 59 years, 60 to 74 years, and patients greater than 75 years with 66 (26.51%), 44 (17.67%), 27 (10.84%), 15 (6.02%) and 11 (4.42%) respectively. The mean age of patients was 28.18 (SD=20.64) years. The main cause of burn was flame burn accounting for 129 (51.81%). It was followed by scald burn, electric burn, chemical burn, and contact burn with 65 (26.10%), 48 (19.28%), 4(1.61%), and 391.20%) respectively. The vast majority of the patients, 227 (91.20%) of the patients had second-degree burns, and 22 (8.80%) of patients suffered from a third-degree burn. The higher percentage of patients suffered from 1-10% total body surface area burn 92 (36.95%). Then, 67 (26.91%), 37 (14.86%), 34 (13.65%), 14 (5.62%), 5(2.01%) had 11%-20%, 21%-30%, 31%-40%, 41%-50% and greater than 50% burn. In that, 0, 12 (17.91%), 15 (40.54%), 23 (67.65%), 7 (50%) and 2 (40%) mortality was seen for 0-10%, 11%-20%, 21%-30%, 31%-40%, 41%-50% and greater than 50% respectively as shown in Table 4.5. The mean %TBSA burn was 19.43 (SD=13.88). The mean length of hospital stay was 10.82 days. The minimum hospital stay was 1 day and the maximum was 97 days. Burn patients with MDR, XDR, and single drug resistance were 88 (61.54%), 36 (61.54%), and 19 (13.29%) respectively.

As shown in Table 1, the total antibiotic consumption among burn patients was 2215.38 measured as DDD. This value corresponds to 82.23 DDD/100BD. The highest DDD/100BD was for ceftriaxone 19.05 (513.26 DDD), followed by cefazolin 15.50 (417.46 DDD) and Piperacillin Tazobactam combination 13.44 (362.07 DDD). The lowest DDD/100 BD was for the combination of ampicillin and cloxacillin with 0.06 DDD/100 BD (1.75DDD). It was preceded by cefepime with 0.12 DDD/100BD (3.20 DDD).

**Table 1.** Defined daily dose (DDD) and DDD/100BD of all antibiotics used in burn patients.

|  |  |  |  |  | % Total |
| --- | --- | --- | --- | --- | --- |
| Class | Antibiotics | ATC code | DDD | DDD/100BD | DDD |
| **Cephalosporin** | Ceftriaxone | J01DD04 | 513.26 | 19.05 | 23.17 |
| **Cephalosporin** | Cefazolin | J01DB04 | 417.46 | 15.50 | 18.84 |
| **Penicillins** | Piperacillin +Tazobactam | J01CR05 | 364.36 | 13.52 | 16.45 |
| **Cephalosporin** | Cefalexin | J01DB01 | 173.06 | 6.42 | 7.81 |
| **Aminoglycoside** | Amikacin | J01GB06 | 167.25 | 6.21 | 7.55 |
| **Fluroquinolone** | Levofloxacin | J01MA12 | 85.50 | 3.17 | 3.86 |
| **Oxazolidinone** | Linezolid | J01XX08 | 70.25 | 2.61 | 3.17 |
| **Macrolides** | Azithromycin | J01FA10 | 67.50 | 2.51 | 3.05 |
| **Tetracycline** | Doxycycline | J01AA02 | 42.00 | 1.56 | 1.90 |
| **Fluroquinolone** | Ofloxacin | J01MA01 | 41.75 | 1.55 | 1.88 |
| **Polymyxins** | Colistin | J01XB01 | 40.51 | 1.50 | 1.83 |
| **Sulfonamides** | Cotrimoxazole | J01EE01 | 40.25 | 1.49 | 1.82 |
|  | AmoxicillinClavulanic |  |  |  |  |
| **Penicillins** | acid* | J01CR02 | 22.80 | 0.85 | 1.03 |
| **Fluroquinolone** | Ciprofloxacin* | J01MA02 | 22.75 | 0.84 | 1.03 |
| **Aminoglycoside** | Gentamicin | J01GB03 | 19.85 | 0.74 | 0.90 |
| **Cephalosporin** | Cefoperazone+Sulbactam | J01DD62 | 19.50 | 0.72 | 0.88 |
| **Cephalosporin** | Ceftazidime | J01DD02 | 15.50 | 0.58 | 0.70 |
| **Cephalosporin** | Cefixime | J01DD08 | 12.60 | 0.47 | 0.57 |
| **Cephalosporin** | Ceftriaxone+Sulbactam | J01DD54 | 11.50 | 0.43 | 0.52 |
| **Imidazole** | Metronidazole | J01XD01 | 11.16 | 0.41 | 0.50 |
| **Carbapenems** | Meropenem | J01DH02 | 8.50 | 0.32 | 0.38 |
| **Penicillins** | Flucloxacillin | J01CF05 | 8.25 | 0.31 | 0.37 |
| **Phenicol** | Chloramphenicol | J01BA01 | 7.00 | 0.26 | 0.32 |
| **Cephalosporin** | Cefotaxime | J01DD01 | 6.54 | 0.24 | 0.30 |
| **Glycopeptide** | Vancomycin | J01XA01 | 6.42 | 0.24 | 0.29 |
| **Penicillins** | Cloxacillin | J01CF02 | 5.63 | 0.21 | 0.25 |
| **Tetracycline** | Tigecycline | J01AA12 | 5.50 | 0.20 | 0.25 |
| **Imidazole** | Metronidazole | P01AB01 | 3.80 | 0.14 | 0.17 |
| **Cephalosporin** | Cefepime | J01DE01 | 3.20 | 0.12 | 0.14 |
| **Penicillins** | Ampicillin+cloxacillin | J01CR50 | 1.75 | 0.06 | 0.08 |
| **Total** |  |  | 2215.38 | 82.23 | 100.00 |

ATC: Anatomic Therapeutic Classification, BD: Bed Days
\* : Two DDD have been defined for oral and parenteral use

Out of 32 antibiotics, there were a total of 12 drugs in the DU90% segment. The first place was for ceftriaxone followed by cefazolin and a combination of piperacillin+tazobactam with 23.32%, 18.96%, and 16.45% respectively as shown in Fig 1.

**Fig 1.**
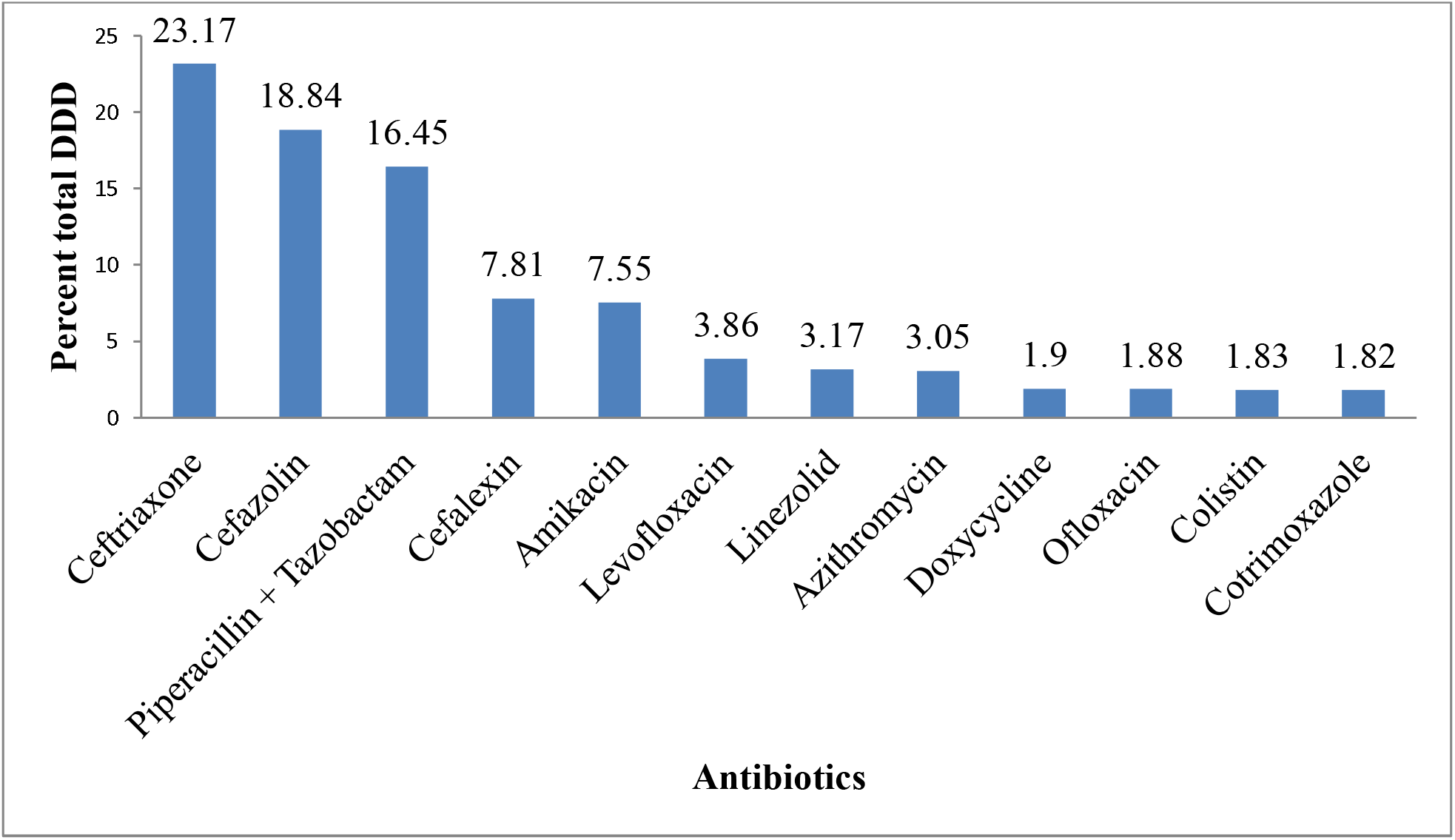
Drug Utilisation 90%.

In the present study, most of the patients 97 (38.96%) were prescribed monotherapy. Following this 85 (34.14%), 33 (13.25%), and 19 (7.63%) of patients have prescribed two, three, and four antibiotics respectively. Only 6.02% of patients have prescribed antibiotics five and above as per Table 2.

**Table 2.** Number of antibiotics prescribed per prescription.

| Number of antibiotics<br>prescribed | Number of patients and percentage (%) (n=249) |
| --- | --- |
| 1 | 97 (39.0) |
| 2 | 85 (34.1) |
| 3 | 33 (13.3) |
| 4 | 19 (7.6) |
| 5 and above | 15 (6.0) |

The mean days of antibiotic therapy were 9.12 (SD=6.21) days. The range of antibiotic therapy was from 1-43 days. In this study, 132 (53.01%) of the patients received antibiotic therapy for 7-14 days, 71 (28.51%), and 35 (14.06%) of patients received antibiotic therapy for 2-6 days and >14 days respectively. Only 11 (4.42%) of patients received antibiotic therapy for 1 day.

The total number of antibiotics prescribed to 249 patients was 2560 and the average number of antibiotics was 2.12 with a range of 1 to 7 prescribed at different points of time.

Out of 249 prescriptions majority of the patients, 137 (58.23%) were given empirical antibiotic therapy. Only 20 (16.06%) of patients were given antibiotic therapy according to the culture report whereas 72 (28.92%) of patient ‘s empirical antibiotic therapy was changed after the review of culture as per Table 3.

**Table 3.**
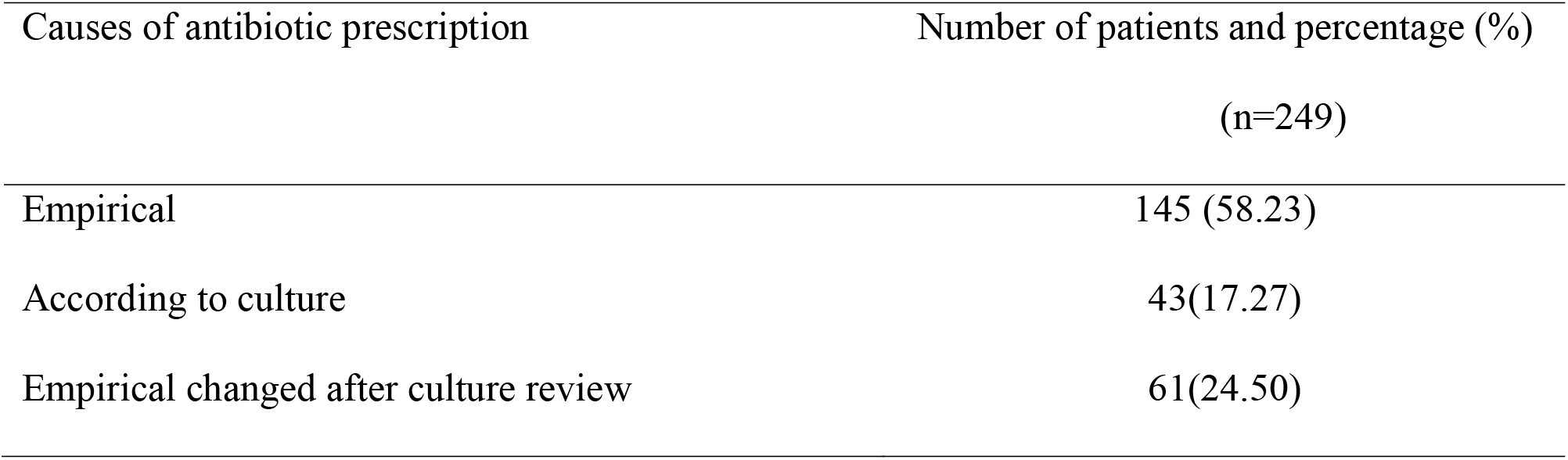
Causes of antibiotic prescription.

## Discussion

Among the study sample, the percentage of female burn patients (51.8%) was more than male burn patients (48.2%). This is similar to the study conducted by Khattak et al. [14] and Dahal et al. [15] This may be due to greater participation of women in household work like cooking and also most of the people are unaware of burn safety measures in Nepal [16]. The main cause of burn was flame burn (51.8%) followed by scald burn (26.1%). This result is consistent with other studies [17, 18]. It could be due to the involvement of females in household activities like cooking [16]. For depth of burn, 91.20% of patients have second degree burn. The result is similar to the study conducted by Ringo et al. [19] and Eser et al. [20] where second-degree burn was higher among burn patients. The variation in depth of burn may depend on the cause of the burn. There is a possibility that the second-degree burn injuries were more common in patients with flame or scalding burn injuries [19] and in this study the main cause of burn was flame and scald burn. The mean TBSA burn was 19.43%. Most of the patients had 0-10% of TBSA burn and the mortality rate was 23.7%. The current study was similar to other studies [21, 22]. The mean length of stay of burn patients in this study was 10.82 days which is consistent with the study done by Amissah et al. [23] and Hop et al. where the mean length of stay was 11 days and 12 days respectively [24].

There are very limited studies done on antibiotic utilization in a burn unit. In this study, the antibiotic utilization pattern for third-generation, ceftriaxone had the highest DDD/100BD with 19.05. This was followed by first-generation cephalosporin, cefazolin (15.50), and penicillin group with the combination of piperacillin and tazobactam (13.52) respectively. Consumption was least for the combination of ampicillin and cloxacillin with 0.06 DDD/100BD. The higher use of ceftriaxone could be due to its broad-spectrum activity. Empirical treatment is usually started when culture is not available in burn patients to reduce the risk of infections. The antibiotic usually chosen for empirical treatment has broad spectrum activity [25]. In contrast, the study conducted by Chauhan et al. showed the highest DDD/100 BD was for cefoperazone, third-generation cephalosporin (40.21 DDD/100 BD) and the least was for second-generation cefuroxime (0.04 DDD/100BD) [26].^26^

Altogether 30 antibiotics were used in this study. This wide variation in the use of antibiotics reflects antibiotic resistance. In the present study, multidrug resistance was seen in 35.3% of burn patients, whereas only 14.5% of patients had extended drug resistance. Another reason for such variation in antibiotics could be due to physician ‘s preference for the use of the wide spectrum of antibiotics. To the best of my knowledge there is no study on DU90% for antibiotics in a burn unit and thus could not be compared based on DU90%. The number of antibiotics which came under DU90% was twelve out of thirty antibiotics.

In the present study, the most commonly prescribed antibiotics were third-generation cephalosporin which is in accordance with the finding of studies done by Chauhan et al. [26] and Padwal et al. [3] but differed in terms of the selection of antibiotics. Ceftriaxone was mostly prescribed in the current study whereas the most prescribed antibiotic was cefoperazone and ceftazidime in the study of Chauhan et al. and Padwal et al. respectively. According to Soleymanzadeh-Moghadam et al. [27] beta-lactam antibiotics were the most frequently prescribed class. The majority of the patients were prescribed cefepime. According to Khairnar et al. [2] in terms of antibiotics, the most commonly used antibiotics were inj gentamicin, inj cefoperazone+sulbactam, inj ciprofloxacin and inj metronidazole. According to Fransen et al.,[28]^28^ most commonly prescribed antibiotics were penicillin (isoxazolylpenicillin), followed by cephalosporins, carbapenems, piperacillin/amoxicillin + beta-lactamase inhibitors and quinolones. This variation may be due to the prevalence different pattern of infecting organisms [29].

The average number of drugs per prescription is an important parameter of a prescription audit. To reduce the risk of drug interactions, the development of bacterial resistance, and hospital costs, the number of drugs per prescription should preferably be as low as possible [30]. We did not look at the co-prescribed drugs here but focused only on antibiotics. In the present study, the average number of antibiotics per prescription was 2.12 with a range of 1 to7. The average number of antibiotics from the present study is comparable with Chauhan et al. [26] and Padwal et al. [3] where an average number of antibiotics prescribed was 2.4 and 2.5 and the range of antibiotics were 1 to 7 and 1 to 4 respectively.

It was observed that the average days of antibiotic therapy were 9.12 days and a majority of the patients received antibiotic therapy for 7-14 days. According to the Guidelines from the French Society for Burn Injuries antibiotic therapy lasting for 7-8 days is recommended [25]. Longer the duration of antibiotic therapy, the greater will be the selection pressure leading to the development of resistant strain which ultimately increases the unnecessary cost of treatment and many a time can cause toxicity in patients [3].

In this study, most of the patients (55.02%) were given empirical therapy. The higher use of empiric antibiotic therapy could be due to the reason that antibiotic therapy should be started immediately. So, it is often started when an infection is suspected and empiric antibiotic therapy is started when bacteriological documentation is lacking. But, this empiric antibiotic therapy may be inappropriate and is known to increase mortality [25]. In a study carried out by Padwal et al. [3] 64% of the patients received antimicrobials prophylactically. Prophylactic use of antibiotics in burn patients is controversial. According to Ramos et al. [31] there is no enough evidence that shows the task of systemic antibiotic prophylaxis in the management of the majority of burn patients. However, the role of systemic antibiotic prophylaxis may come into existence if it is used in patients with severe burns who require mechanical ventilation, and in selected split-thickness skin grafting procedures.

The limitations of this study were: 1) Data were collected manually as there was no digital system to know the record of the patient ‘s details. So there may be the chance of missing data. 2) Since the study was retrospective, it was difficult to quantify the number of topical antibiotics and DDD was not assigned for it. For this reason, topical antibiotics were not included in this study.

## Conclusion

This study showed the general trend in antibiotic utilization patterns in burn patients. Third-generation cephalosporin, ceftriaxone was the most prescribed antibiotic. Since antibiotics are the key drugs used for the treatment of burn patients, a similar future prospective is needed to conduct regular antibiotic consumption using the DDD methodology for regular monitoring of antibiotic consumption so that timely intervention can be made and this study results can be used as a baseline study. Moreover, electronic patient records can be introduced to obtain more precise information on the actual situation with antibiotic consumption.

## Data Availability

All data produced in the present work are contained in the manuscript.

## Acknowledgement

I am thankful to WHO collaborating centre for Drug Statistics Methodology, Oslo, Norway and especially to Hege Salvesen Blix for answering my queries regarding use of DDD as a method of drug consumption.

